# 8,266 SARS-CoV-2 Genomic Assemblies from Asymptomatic Carriers in Japan

**DOI:** 10.1101/2025.07.29.25332287

**Authors:** Hajime Ohyanagi, Junko S Takeuchi, Yuichi Kawanishi, Saya Ohi, Teiichiro Shiino, Moto Kimura, Yusuke Takahashi, Shigeru Yoshida, Minoru Kato, Yukumasa Kazuyama, Masato Ikeda, Wataru Sugiura

## Abstract

In the context of public health, asymptomatic carriers of respiratory infectious diseases are considered a hidden yet critical factor in the transmission of infection and represent a key target for efforts to mitigate disease spread. To contribute a unique genomic resource to the SARS-CoV-2 research community, we collected SARS-CoV-2–positive samples from asymptomatic individuals at the SB Coronavirus Inspection Center Corp. during the COVID-19 pandemic in Japan and conducted a comprehensive analysis of their viral genomes. Using Illumina COVIDSeq technology, we successfully generated 8,266 SARS-CoV-2 genome assemblies, all of which have been made publicly available to facilitate further research. In this report, we summarize our efforts to collect SARS-CoV-2–positive samples from asymptomatic individuals and highlight the key features and accessibility of this genomic dataset.

## Background & Summary

We operate the SB Coronavirus Inspection Center Corp. (SBCVIC) ^1^, a large-scale screening facility specializing in SARS-CoV-2 testing for individuals without symptoms. The center primarily serves workplace-based routine screening programs and voluntary screening requests from local governments, and therefore most samples originated from individuals who did not report symptoms at the time of testing. Epidemiologically, asymptomatic carriers of SARS-CoV-2 are known to have non-negligible transmission capabilities comparable to those of symptomatic individuals^2–5^. Therefore, their existence represents a hidden yet critical factor in controlling the spread of the pandemic^6,7^. From a genomic perspective, samples from asymptomatic carriers are also a valuable resource for investigating genetic factors in the viral genome that may be associated with the presence or absence of clinical symptoms in COVID-19 patients^8^.

During the pandemic period in Japan^9,10^, spanning from July 2020 to January 2023, the country experienced eight epidemic waves of SARS-CoV-2 infection^11^ (**Figure 1A)**. These included waves dominated by the Alpha (the 4^th^ wave), Delta (the 5^th^ wave), and Omicron sublineages BA.1, BA.2, and BA.5 (from the 6^th^ wave onward) (**Figure 1B** for symptomatic cases, and **Figure 1C** for asymptomatic cases). Over the course of the study (July 27, 2020 – January 16, 2023), we tested a total of 4,573,575 samples, among which 18,475 tested positive for SARS-CoV-2 using reverse transcription real-time quantitative polymerase chain reaction (RT-qPCR) (Ct values of ≤ 40) (**Figure 2** and **Table 1**). The overall average positive rate among asymptomatic individuals was 0.40%. The positive rates varied by wave: 0.05% during the 3^rd^ wave, 0.13% in the 4^th^ wave (Alpha), 0.23% in the 5^th^ wave (Delta), 4.5% in the 6^th^ wave, 8.2% in the 7^th^ wave (BA.1/BA.2), and 5.2% in the 8^th^ wave (BA.5). The number of newly confirmed cases in Japan (**Figure 1A**) shows a trend similar to the positive rate among asymptomatic individuals mentioned above.

**Figure 1.**
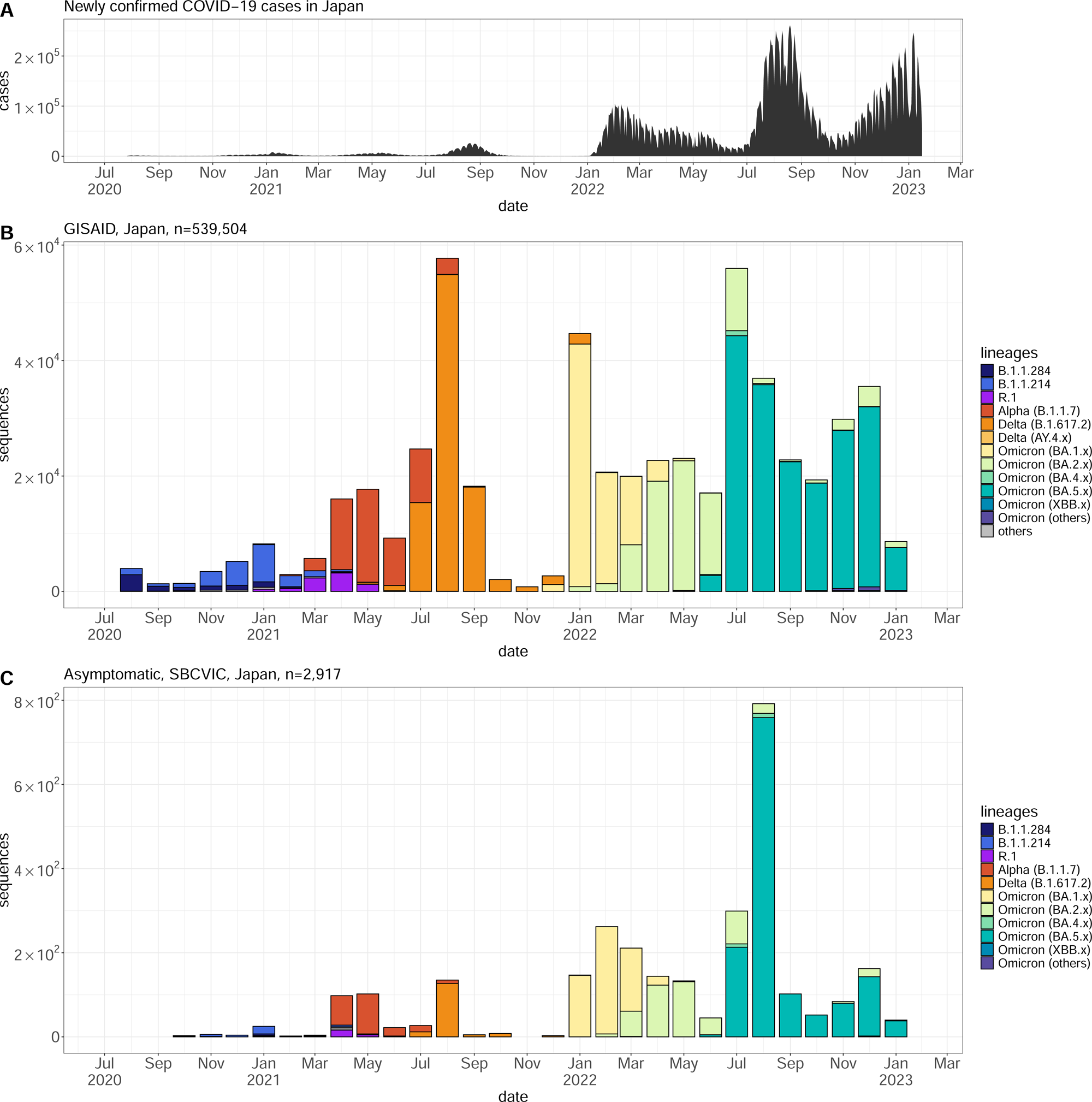
Monthly epidemiological distribution of SARS-CoV-2 variants in Japan (July 27, 2020 - January 16, 2023). (A) Monthly number of newly confirmed COVID-19 cases in Japan, based on open data from the Ministry of Health, Labour and Welfare (accessed on May 1, 2023). (B) All domestic sequences registered in the Global Initiative on Sharing All Influenza Data (GISAID) EpiCoV database as of February 6, 2023 (n = 539,504). (C) Positive cases detected in asymptomatic individuals in this study (n = 2,917).

**Figure 2.**
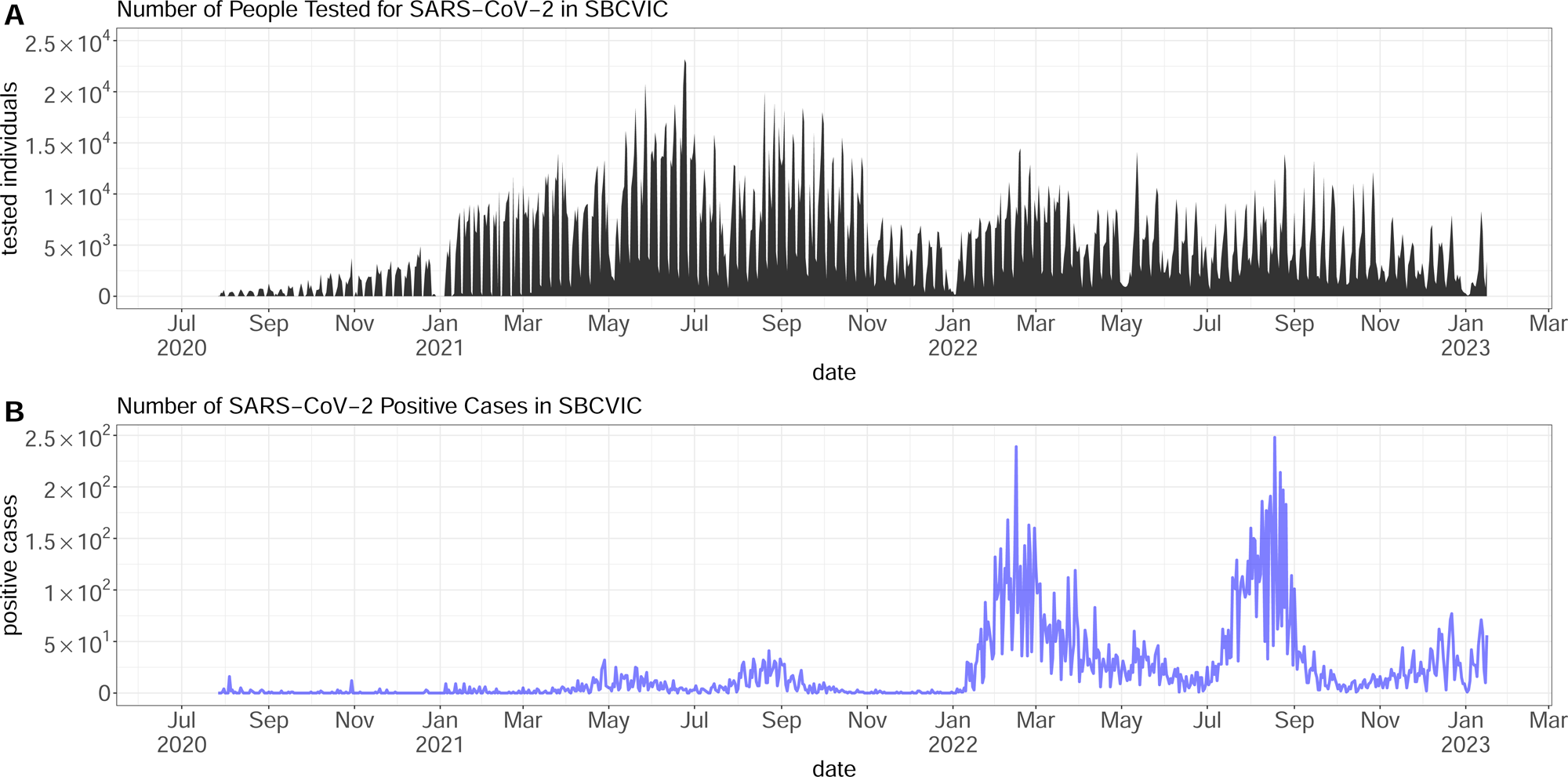
Number of tested (gray) and SARS-CoV-2-positive (blue) cases in this study. Between July 27, 2020, and January 16, 2023, a total of 4,573,575 saliva samples were tested (A), of which 18,475 were positive by RT-qPCR testing (B). Individuals who opted out of the study were excluded from this count.

**Table 1.** Statistics of the study participants. . Gender is shown in n (%), and age is presented as median [interquartile range].

|  | Value |
| --- | --- |
| **Tested individuals** | 4,573,575 |
| **Positive cases** | 18,475 |
| **Gender** |  |
| **Male** | 6,408 (34.7%) |
| **Female** | 5,944 (32.2%) |
| **Unknown** | 6,123 (33.1%) |
| **Age (Years)** | 36 [27 - 44] |

Starting October 9, 2020 (approximately six weeks after the initiation of the study), we began collecting background information on positive cases from 45 prefectures across Japan, including Shiga, Tokyo, Nagasaki, Osaka, Hokkaido, Fukuoka, Chiba, Gifu, Kanagawa, Saitama, Hyogo, Aichi, Kyoto, Miyagi, Gumma, Hiroshima, Nagano, Kumamoto, Ibaraki, Shizuoka, Tochigi, Fukushima, Yamanashi, Nara, Miyazaki, Okinawa, Mie, Ehime, Iwate, Okayama, Kagawa, Yamagata, Saga, Aomori, Akita, Niigata, Toyama, Tottori, Ishikawa, Yamaguchi, Oita, Kagoshima, Fukui, Shimane, and Kochi (**Figure 3**). No positive case was collected from two prefectures (Tokushima and Wakayama). Among 18,475 positive cases, 6,408 (34.7%) were male, 5,944 (32.2%) were female, and 6,123 (33.1%) had missing gender information. Age information was available for 12,544 individuals, among whom the median age was 36 years (interquartile range [IQR]: 27–44). (**Table 1**).

**Figure 3.**
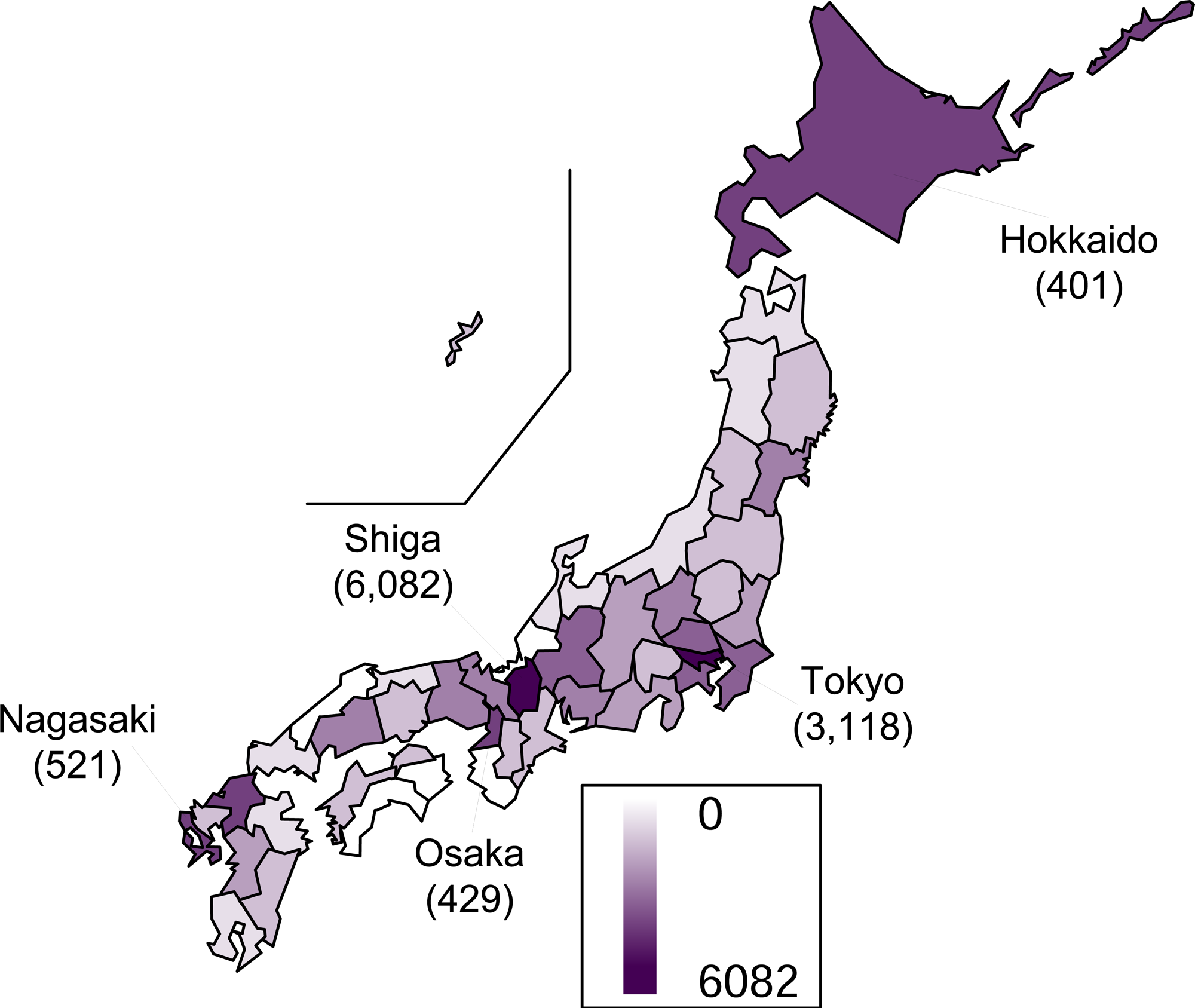
Geographic distribution of sample collection. Color intensity indicates the number of samples collected, with darker shades representing higher counts. The top five prefectures with the highest number of samples were Shiga (n = 6,082), Tokyo (n = 3,118), Nagasaki (n = 521), Osaka (n = 429), and Hokkaido (n = 401).

In total, we obtained 18,475 SARS-CoV-2-positive samples from asymptomatic individuals. We performed whole-genome sequencing on residual saliva samples and assembled viral genome sequences that met quality thresholds for downstream analysis. Ultimately, we successfully generated 8,266 genome assemblies from these cases, which are archived in the SBCVIC genomic dataset and categorized into two groups (**Table 2** and **Table 3**) according to operational criteria (based on genome length and Ct values).

**Table 2.**
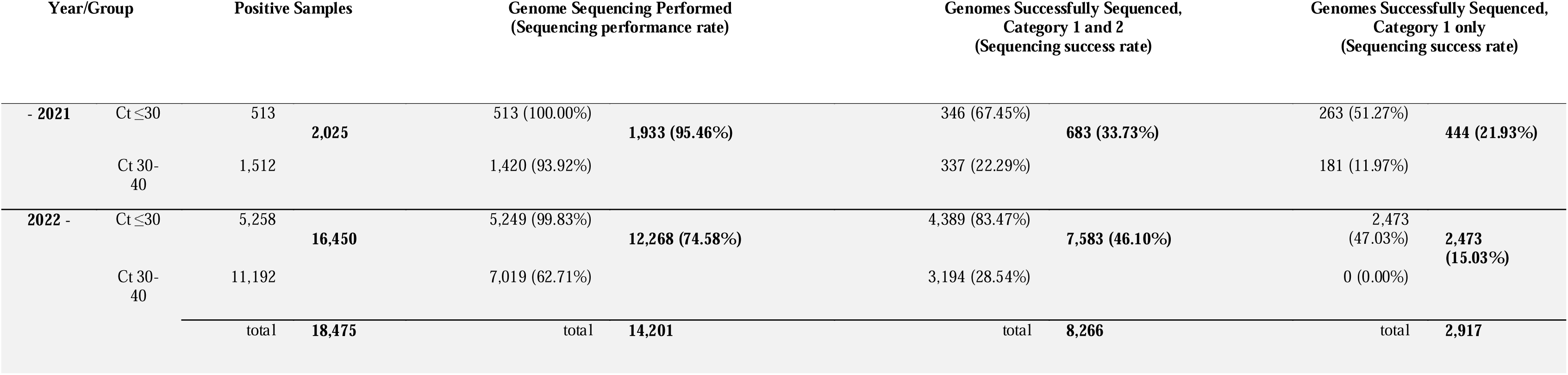
Summary of viral genome sequencing. The positive samples (18,475 samples in total) were selectively implemented for genome sequencing (14,201 samples in total). Eventually, 8,266 samples were successfully sequenced in both quality categories (Category 1 and Category 2), while Category 1 consists of 2,917 samples. For the definition of Categories 1 and 2, see Table 3.

**Table 3.** Submission of genomic sequences to public repositories (INSDC/DDBJ and GISAID). In total, 8,266 genomic assemblies were submitted to the public repositories in this dataset. * BioSample accessions provide metadata describing the biological source materials used to generate sequencing data. ** 219 BioSample data did not meet Category 2 criteria; sequence data for these were not submitted.

| Category and Criteria | Number of Sequences | BioSample* Accession | Sequence Accession | SRA Run Accession | SRA Experiment Accession | GISAID ID |
| --- | --- | --- | --- | --- | --- | --- |
| <u>Category 1:</u><br>Genome length ≥ 29,000 nucleotides and<br>Ct value ≤ 40 (for - 2021)<br>≤30 (for 2022 -) | 2917 | SAMD00738313<br>- SAMD00741057,<br>SAMD00741082,<br>SAMD00786981<br>- SAMD00787151 | BS007837<br>- BS010581,<br>BS010689,<br>BS016048<br>- BS016218 | DRR600457<br>- DRR603201,<br>DRR603222,<br>DRR608552<br>- DRR608722 | DRX581004<br>- DRX583748,<br>DRX583769,<br>DRX589099<br>- DRX589269 | EPI_ISL_19576276<br>- EPI_ISL_19582591,<br>EPI_ISL_19582637,<br>EPI_ISL_19575435<br>- EPI_ISL_19575605 |
| <u>Category 2:</u><br>Complete Genome not meeting<br>Category 1 criteria<br>and<br>with unidentified nucleotides ≤ 20% | 5349 | SAMD00741058<br>- SAMD00741081,<br>SAMD00741083<br>- SAMD00746626** | BS010669<br>- BS010688,<br>BS010690<br>- BS016018 | DRR603202<br>- DRR603221,<br>DRR603223<br>- DRR608551 | DRX583749<br>- DRX583768,<br>DRX583770<br>- DRX589098 | EPI_ISL_19582592<br>- EPI_ISL_19582606,<br>EPI_ISL_19582632<br>- EPI_ISL_19582636,<br>EPI_ISL_19582638<br>- EPI_ISL_19585391,<br>EPI_ISL_19579847<br>- EPI_ISL_19580766,<br>EPI_ISL_19578927<br>- EPI_ISL_19579846,<br>EPI_ISL_19574700<br>- EPI_ISL_19575434 |

For technical validation, we evaluated the distribution of consensus genome sequence lengths and examined the lineage composition of the SARS-CoV-2 genomes. We also assessed the presence of known mutations to confirm the reliability of our sequencing pipeline for allele typing. In particular, we focused on two adjacent, co-occurring mutations in the nucleocapsid (N) protein R203K/G204R^12^, which are associated with increased viral infectivity, fitness, and virulence. For further details, see the **Technical Validation** section.

## Methods

### Participants

Procedures complied with national, institutional committees on human experimentation and with the Helsinki Declaration (1975, revised in 2008). No participants were directly recruited for this study. All analyses were performed using saliva specimens collected by SBCVIC testing laboratories as part of their routine SARS-CoV-2 testing for asymptomatic individuals. In Japan, explicit opt-in informed consent is not required for this type of specimen. The institutional observational studies employed an IRB-approved opt-out process in accordance with national ethical guidelines. Study information and opt-out procedures were publicly posted on the institutional websites, and individuals were given the opportunity to decline the secondary use of their specimens and associated clinical information. Saliva specimens collected by the testing laboratories were handled under their approved protocol, only anonymized samples and limited clinical data were provided to our research team. The study protocol was approved by the Institutional Review Board of the National Center for Global Health and Medicine (approval number: NCGM-G-004225-00, August 5, 2020).

### SARS-CoV-2 Testing (Screening of Asymptomatic Cases)

Following previously described protocols^1,13^, we conducted SARS-CoV-2 testing and genome sequencing. The SBCVIC provided routine workplace-based SARS-CoV-2 screenings upon company request, as well as voluntary screenings requested by local governments. Each participant self-collected approximately 2 mL of saliva using the ZEESAN Saliva RNA Sample Collection Kit (MD-ZSV-001; Zeesan Biotech, Fujian, China), which contained 1 mL of a guanidine-based viral inactivation buffer. The sample was classified and shipped as UN 3373 Biological Substance, Category B. On the day of arrival, SARS-CoV-2 RT-qPCR testing was performed using the SARS-CoV-2 Direct Detection RT-qPCR Kit (Takara Bio, Shiga, Japan). Kits RC30JW and RD003 were used until April 4 and from April 5, 2021, respectively, according to the manufacturer’s protocol. Because the guanidine-based viral inactivation buffer can inhibit RT-PCR reactions^14^, we modified the pretreatment process. Specifically, 16 µL of the saliva/buffer mixture was combined with 16 µL of water and 4 µL of pretreatment reagent (Solution A, included in the kit) to dilute the inhibitor. The resulting 36 µL mixture was incubated at room temperature for 5 minutes, followed by 5 minutes at 95°C. A 2.5 µL aliquot of this treated mixture was used as the RT-qPCR template. Test results (positive or negative) were determined based on the protocol, with Ct values ≤ 40 considered positive.

### Viral Genome Sequencing

Samples from SARS-CoV-2-positive individuals were subjected to genome sequencing. For RNA extraction, 50 μL of viral nucleic acid was extracted from 200 μL of saliva samples using MagMAX Viral/Pathogen Nucleic Acid Isolation Kit (Thermo Fisher Scientific, MA, USA) with MVP_Saliva_200_Flex_V1 protocol for KingFisher Flex System (Thermo Fisher Scientific, MA, USA). The eluted RNA (20 μL) was treated with RQ1 RNase-Free DNase (Promega, WI, USA). Subsequent cDNA synthesis, target amplification using the ARTIC^15^ primer set (versions used for each sample are documented in the BioSample archive), and library preparation were performed according to the Illumina COVIDseq Test Reference Guide (Illumina Inc., CA, USA). The sequencing performance rates were 100.00% (-2021) and 99.83% (2022-) for samples with Ct values ≤ 30 across all periods (see the column **Genome Sequencing Performed** in **Table 2**). However, the rate decreased for higher Ct values (30–40), particularly from 2022 onward, due to operational constraints. Sequencing was conducted on an Illumina NextSeq 2000 system.

### Bioinformatics

Sequencing data were analyzed using the Illumina DRAGEN COVID Lineage Pipeline (https://jp.illumina.com/products/by-type/informatics-products/basespace-sequence-hub/apps/dragen-covid-lineage.html). Consensus sequences in FASTA format were generated (see **Data Records** and **Data Overview**). The DRAGEN versions used for each sample are documented in the BioSample archive. Sequences equal to or longer than 29,000 nucleotides in length (Category 1 in **Table 2** and **Table 3**) were analyzed for lineage assignment using Phylogenetic Assignment of Named Global Outbreak Lineages (PANGOLIN) version 4.3.1 (pangolin-data 1.25.1) ^16^.

For variant detection, the reference genome used was the Wuhan WIV04 strain (hCoV-19/Wuhan/WIV04/2019, EPI_ISL_402124), downloaded from GISAID^17–19^. A multiple sequence alignment, including the WIV04 reference, was generated in FASTA format using MAFFT (online version 7) ^20^. The WIV04 sequence was then removed from the alignment, which was analyzed using AliView (version 1.28) ^21^ and custom Python scripts (Python version 3.12.3^22^) to calculate allele frequencies at the R203K/G204R sites in the N protein. Python scripts were also used to perform statistical test (G-test).

To analyze the distribution of SARS-CoV-2 lineages across Japan during the study period (July 27, 2020 – January 16, 2023), we utilized sequences registered in the GISAID EpiCov database (https://gisaid.org) as of February 6, 2023. The filtering criteria were:

- Host: Human
- Location: Asia / Japan
- Collection date: July 27, 2020 – January 16, 2023
- Sequence length: ≥ 29,000 nt
- Passage history: Original

The resulting 539,504 sequences were analyzed using PANGOLIN for lineage classification (see **Figure 1B**).

Data visualization was performed using R version 4.4.0^23^ and the R package *NipponMap*^24^.

### Data Records

This dataset includes:

- Raw sequencing reads (SRA accessions under DRP012098)
- Consensus genome assemblies (FASTA format)
- BioSample metadata (BioSample accessions listed in the **Data Overview** section)
- Lineage annotations (PANGOLIN)

All files follow International Nucleotide Sequence Database Collaboration (INSDC)^25^-compliant formats. The PANGOLIN lineage annotations are archived exclusively in GISAID and are not part of the INSDC repositories.

### Data Overview

Of the 18,475 SARS-CoV-2-positive samples, 14,201 were subjected to viral genome sequencing. By the end of December 2021, sequencing was implemented for 100% of samples with Ct ≤30 and 93.92% of those with Ct value between 30–40. From 2022 onward, the coverage remained high Ct ≤30 samples (for 99.83%) but decreased to 62.71% for Ct 30–40 samples (**Table 2**). Across all sequencing attempts, a total of 8,266 SARS-CoV-2 genomes were successfully assembled. Among these, 2,917 sequences met the criteria for Category 1, and the remaining genomes were classified as Category 2, as defined in **Table 3**. These final yields reflect both Ct value–dependent feasibility and operational constraints during the study period.

Raw reads and consensus genome sequences for 8,266 SARS-CoV-2 samples were submitted and are available for download from the NCBI Sequence Read Archive in the International Nucleotide Sequence Database Collaboration (INSDC)^25^, under the study accession number DRP012098^26^. While 1,248 of them were used in previous publication^1^, all high-quality genome assemblies are currently available under the single accession. The corresponding BioSample and other accession numbers are summarized in **Table 3**. According to operational criteria (based on genome length and Ct values), sequences were categorized into two groups (see **Table 3)**. Assembled genome sequences were also submitted to GISAID, under the accession IDs listed in **Table 3**.

### Technical Validation

#### Statistics of Consensus Genome Sequences

We first examined the length distribution of the 8,266 complete consensus genome sequences across both Category 1 and Category 2 (**Figure 4**). A sufficient number (n = 2,917) of nearly full-length genome sequences (≥ 29,000 nt; Category 1) were included and exclusively used for SARS-CoV-2 lineage assignment via PANGOLIN, as well as for subsequent analyses described below.

**Figure 4.**
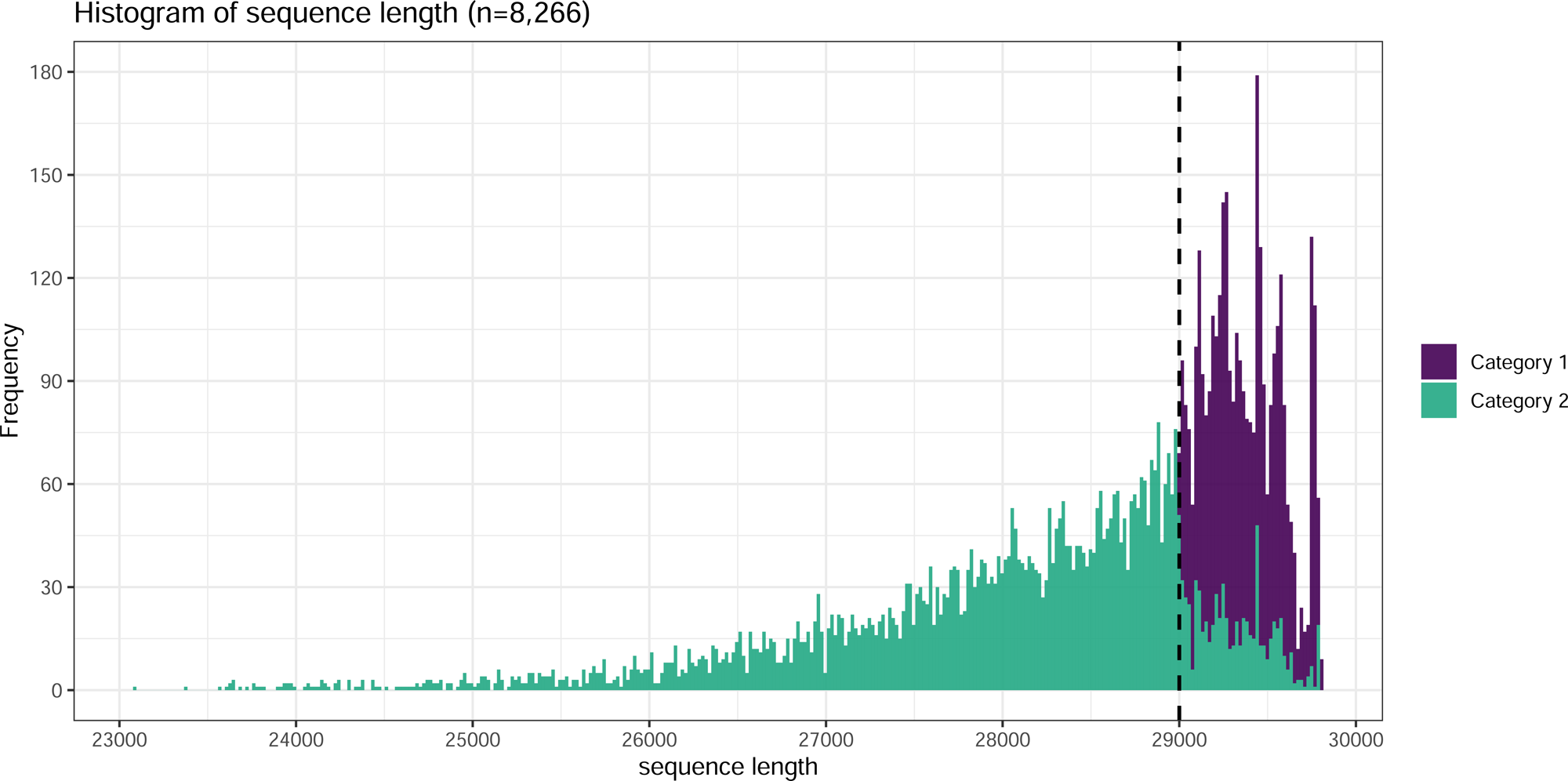
Distribution of SARS-CoV-2 genome consensus sequence lengths obtained in this study (n = 8,266). A histogram shows the distribution of sequence lengths (X-axis) and their frequencies (Y-axis). Category 1 sequences (n = 2,917) are shown in dark purple, and Category 2 sequences (n = 5,349) are shown in green. The dashed line represents the threshold for near-complete genome sequences (29,000 nt).

#### Epidemiological Distribution of SARS-CoV-2 Variants

Since the SBCVIC dataset represents a SARS-CoV-2 subpopulation within the broader Japanese population, we anticipated a comparable lineage distribution between our dataset and that of GISAID. Lineage analysis using PANGOLIN (**Figure 1B** and **1C**) revealed a similar distribution of SARS-CoV-2 variants between SBCVIC and GISAID datasets, supporting the validity of the overall SBCVIC sampling, sequencing, and analysis workflow.

#### Detection of R203K/G204R Mutations in the Nucleocapsid Protein

The 2,917 Category 1 genome assemblies were also used for further technical validation. As control datasets, we randomly selected and created three replicates of 2,917 SARS-CoV-2 genome sequences from symptomatic individuals of Japanese origin deposited in GISAID during the same period (July 27, 2020 to January 16, 2023) (**Table 4**). To demonstrate the genotyping capability of our dataset, we focused on two well-known adjacent co-occurring mutations R203K/G204R in the N protein of SARS-CoV-2, which are known to affect viral pathogenicity^12^.

**Table 4.** Number of wild-type and mutant alleles at the nucleocapsid protein mutation sites (R203K/G204R) in symptomatic (GISAID, three replicates) and asymptomatic (SBCVIC, this study) populations.

|  | Wild allele<br>(R203/G204) | Mutant allele<br>(203K/204R) | Allele Not<br>Determined |
| --- | --- | --- | --- |
| **GISAID (Symptomatic, replicate 1, n=2917)** | 7 | 2391 | 519 (17.8%) |
| **GISAID (Symptomatic, replicate 2, n=2917)** | 2 | 2365 | 550 (18.9%) |
| **GISAID (Symptomatic, replicate 3, n=2917)** | 7 | 2365 | 545 (18.7%) |
| **SBCVIC (Asymptomatic, n=2917)** | 7 | 2739 | 171 (5.9%) |

Genome sequences were aligned to the SARS-CoV-2 reference genome by MAFFT, and the allele frequencies at the R203K/G204R sites were calculated for both the GISAID (symptomatic) and SBCVIC (asymptomatic) groups (see **Methods**). Our SBCVIC dataset exhibited a lower rate of missing allele calls (5.9%) compared to the GISAID datasets (17.8% for replicate 1, 18.9% for replicate 2, and 18.7% for replicate 3), indicating higher call rate across the majority of samples while not showing the higher genotyping quality itself. Interestingly, the frequency of the mutant allele in the SBCVIC (asymptomatic) group was comparable to that in the GISAID (symptomatic) group (**Table 4**) (*P* = 0.799676 for replicate 1, *P* = 0.133389 for replicate 2, and *P* = 0.783957 for replicate 3, G-test), despite our initial expectation that mutation rates would differ between symptomatic and asymptomatic individuals. While we do not propose a specific hypothesis here, this allele-level analysis highlights a potential application of the SBCVIC dataset.

### Usage Note

The dataset of SARS-CoV-2 samples collected and analyzed from asymptomatic individuals in Japan is highly distinctive. We demonstrated the validity of this SBCVIC dataset for analyzing the epidemiological distribution of SARS-CoV-2 variants and for conducting allele-level genotyping. Possible applications of our dataset are, analysis for asymptomatic-specific mutations that would be keys for SARS-CoV-2 virulence (see **Technical Validation**), training AI classifiers and Genome Large Language Models on asymptomatic variant data, and building viral fitness model for SARS-CoV-2. We believe that this dataset has significant potential for epidemiologists, geneticists, and computational biologists to provide insights into the modulation of SARS-CoV-2 virulence and to enhance our understanding of viral fitness.

## Data Availability

All custom scripts used in this study are available on GitHub at https://github.com/oyasai55/8266_SARS-CoV-2_Genomic_Assemblies_Scripts/.

## Acknowledgements

We thank Mr. Takahiro Sakurai and Mr. Hiroshi Ikawa for their support with bioinformatics analyses. We also acknowledge Mr. Tsuyoshi Saito, Ms. Natsumi Miyazaki, Mr. Atsushi Sugi, and Mr. Masanori Tamanaha for their assistance in managing sample metadata.

## Author contributions

H.O. and J.S.T. designed and performed the analyses and wrote the manuscript. J.S.T. and Yuichi K. managed the genome sequence data from SBCVIC. S.O. conducted bioinformatics and statistical analyses. J.S.T. and S.O. conducted figure/table visualization. T.S. and W.S. supervised the entire analysis process. Moto K. directed the research contract with the SBCVIC and intellectual properly. Y.T., S.Y., Minoru K. and Yukumasa K. was responsible for RT-qPCR testing with the viral genome sequencing at SBCVIC. M.I. and W.S. conceived an inspection system by SBCVIC and acquired research funding. All authors critically reviewed and approved the final version of the manuscript.

## References

1 Shiino, T. et al. Molecular epidemiology of SARS-CoV-2 genome sentinel surveillance in commercial COVID-19 testing sites targeting asymptomatic individuals during Japan’s seventh epidemic wave. Sci Rep 14, 20950 (2024). 10.1038/s41598-024-71953-8

2 Buitrago-Garcia, D. et al. Occurrence and transmission potential of asymptomatic and presymptomatic SARS-CoV-2 infections: A living systematic review and meta-analysis. PLoS Med 17, e1003346 (2020). 10.1371/journal.pmed.1003346

3 Buitrago-Garcia, D. et al. Occurrence and transmission potential of asymptomatic and presymptomatic SARS-CoV-2 infections: Update of a living systematic review and meta-analysis. PLoS Med 19, e1003987 (2022). 10.1371/journal.pmed.1003987

4 Casey-Bryars, M., et al. Presymptomatic transmission of SARS-CoV-2 infection: a secondary analysis using published data. BMJ Open 11, e041240 (2021). 10.1136/bmjopen-2020-041240

5 Yang, Q. et al. Just 2% of SARS-CoV-2-positive individuals carry 90% of the virus circulating in communities. Proc Natl Acad Sci U S A 118 (2021). 10.1073/pnas.2104547118

6 Davis, J. T. et al. Cryptic transmission of SARS-CoV-2 and the first COVID-19 wave. Nature 600, 127–132 (2021). 10.1038/s41586-021-04130-w

7 Nabeshima, T. et al. COVID-19 cryptic transmission and genetic information blackouts: Need for effective surveillance policy to better understand disease burden. Lancet Reg Health West Pac 7, 100104 (2021). 10.1016/j.lanwpc.2021.100104

8 Ferdinand, A. S. et al. An implementation science approach to evaluating pathogen whole genome sequencing in public health. Genome Med 13, 121 (2021). 10.1186/s13073-021-00934-7

9 Kitahara, K., Nishikawa, Y., Yokoyama, H., Kikuchi, Y. & Sakoi, M. An overview of the reclassification of COVID-19 of the Infectious Diseases Control Law in Japan. Glob Health Med 5, 70–74 (2023). 10.35772/ghm.2023.01023

10 Takeuchi, J. S., et al. Large-scale screening of SARS-CoV-2 variants in Tokyo, Japan: A 3-year and 9-month longitudinal survey. Glob Health Med 7, 151–160 (2025). 10.35772/ghm.2025.01004

11 The Ministry of Health, Labour and Welfare, Japan Infectious Disease Trend Visualization System, <https://covid19.mhlw.go.jp/en/> (2023).

12 Wu, H. et al. Nucleocapsid mutations R203K/G204R increase the infectivity, fitness, and virulence of SARS-CoV-2. Cell Host Microbe 29, 1788–1801 e1786 (2021). 10.1016/j.chom.2021.11.005

13 Terada-Hirashima, J. et al. Investigation of the use of PCR testing prior to ship boarding to prevent the spread of SARS-CoV-2 from urban areas to less-populated remote islands. Glob Health Med 4, 174–179 (2022). 10.35772/ghm.2022.01008

14 Katsuno, T. et al. Diagnostic accuracy of direct reverse transcription-polymerase chain reaction using guanidine-based and guanidine-free inactivators for SARS-CoV-2 detection in saliva samples. J Virol Methods 326, 114909 (2024). 10.1016/j.jviromet.2024.114909

15 ARTIC Network, SARS-CoV-2 (COVID-19). https://artic.network/viruses/sars-cov-2.

16 O’Toole, A. et al. Assignment of epidemiological lineages in an emerging pandemic using the pangolin tool. Virus Evol 7, veab064 (2021). 10.1093/ve/veab064

17 Elbe, S. & Buckland-Merrett, G. Data, disease and diplomacy: GISAID’s innovative contribution to global health. Glob Chall 1, 33–46 (2017). 10.1002/gch2.1018

18 Khare, S. et al. GISAID’s Role in Pandemic Response. China CDC Wkly 3, 1049–1051 (2021). 10.46234/ccdcw2021.255

19 Shu, Y. & McCauley, J. GISAID: Global initiative on sharing all influenza data - from vision to reality. Euro Surveill 22 (2017). 10.2807/1560-7917.ES.2017.22.13.30494

20 Katoh, K. & Standley, D. M. MAFFT multiple sequence alignment software version 7: improvements in performance and usability. Mol Biol Evol 30, 772–780 (2013). 10.1093/molbev/mst010

21 Larsson, A. AliView: a fast and lightweight alignment viewer and editor for large datasets. Bioinformatics 30, 3276–3278 (2014). 10.1093/bioinformatics/btu531

22 Python Software Foundation. Python Language Reference, version 3.12.3. https://www.python.org. (2024).

23 R: A Language and Environment for Statistical Computing. R Foundation for Statistical Computing, Vienna, Austria. https://www.R-project.org/. (2024).

24 Tanimura, S. NipponMap: Japanese Map Data and Functions. R package version 0.2, https://cran.r-project.org/web/packages/NipponMap. (2017).

25 Karsch-Mizrachi, I. et al. The international nucleotide sequence database collaboration (INSDC): enhancing global participation. Nucleic Acids Res (2024). 10.1093/nar/gkae1058

26 NCBI Sequence Read Archive, <https://identifiers.org/ncbi/insdc.sra:DRP012098> (2024).

